# Evolution of the Umbilical Cord Blood Proteome Across Gestational Development

**DOI:** 10.1101/2024.06.21.24309280

**Authors:** Leena B. Mithal, Nicola Lancki, Ted Ling-Hu, Young Ah Goo, Sebastian Otero, Nathaniel J. Rhodes, Byoung-Kyu Cho, William A. Grobman, Judd F. Hultquist, Denise Scholtens, Karen G. Mestan, Patrick C. Seed

**Affiliations:** Department of Pediatrics, Division of Infectious Diseases, Ann & Robert H. Lurie Children’s Hospital of Chicago, Northwestern University Feinberg School of Medicine, Chicago, IL, USA; Department of Preventive Medicine, Division of Biostatistics, Northwestern University Feinberg School of Medicine, Chicago, IL, USA; Department of Medicine, Division of Infectious Diseases, Northwestern University Feinberg School of Medicine, Chicago, IL, USA; Center for Pathogen Genomics and Microbial Evolution, Havey Institute for Global Health, Northwestern University Feinberg School of Medicine, Chicago, IL, USA; Mass Spectrometry Technology Access Center at McDonnell Genome Institute (MTAC@MGI), Washington University in Saint Louis School of Medicine, MO, USA; Department of Pharmacy Practice, Midwestern University, College of Pharmacy, Downers Grove, IL, USA; Pharmacometrics Center of Excellence, Midwestern University, Downers Grove, IL, USA; Department of Pharmacy, Northwestern Memorial Hospital, Chicago, IL, USA; Department of Obstetrics and Gynecology, Ohio State University, Columbus, OH, USA; Department of Pediatrics, Division of Neonatology, University of California San Diego, CA, USA

**Author notes:** **Correspondence:** Leena B. Mithal, MD MSCI.

**Keywords:** cord blood, proteomics, neonatal immunology, prematurity, immune development, biomarker development

## Abstract

Neonatal health is dependent on early risk stratification, diagnosis, and timely management of potentially devastating conditions, particularly in the setting of prematurity. Many of these conditions are poorly predicted in real-time by clinical data and current diagnostics. Umbilical cord blood may represent a novel source of molecular signatures that provides a window into the state of the fetus at birth. In this study, we comprehensively characterized the cord blood proteome of infants born between 24 to 42 weeks using untargeted mass spectrometry and functional enrichment analysis. We determined that the cord blood proteome at birth varies significantly across gestational development. Proteins that function in structural development and growth (e.g., extracellular matrix organization, lipid particle remodeling, and blood vessel development) are more abundant earlier in gestation. In later gestations, proteins with increased abundance are in immune response and inflammatory pathways, including complements and calcium-binding proteins. Furthermore, these data contribute to the knowledge of the physiologic state of neonates across gestational age, which is crucial to understand as we strive to best support postnatal development in preterm infants, determine mechanisms of pathology causing adverse health outcomes, and develop cord blood biomarkers to help tailor our diagnosis and therapeutics for critical neonatal conditions.

## Introduction

Neonatal health is dependent on early risk stratification, diagnosis, and timely management of many potentially devastating conditions. Preterm infants are at increased risk of prematurity-related complications, including: early-onset sepsis, chronic lung disease, intraventricular hemorrhage, necrotizing enterocolitis, and neurodevelopmental impairment.^1–3^ Many of these conditions are poorly predicted in real-time by clinical data, including currently available diagnostic testing. Thus, biomarkers have been sought to aid early and targeted treatment and prognosis for these conditions.

Umbilical cord blood may represent a novel source of molecular signatures that provides a window into the state of the fetus at birth. Umbilical cord blood inflammatory markers have been studied as diagnostic indicators of early-onset sepsis^4–6^. Specific cord blood cytokines have been identified as predictors or correlates of retinopathy of prematurity^7^, atopic disease^8^, infantile hemangioma^9^, placental histopathology^10^, and more^4^. However, few of these cord blood biomarkers have been translated into diagnostic tools in clinical practice.

“Omics” methodologies have been previously used to profile amniotic fluid and infant blood to predict pre-eclampsia, preterm birth, and late-onset sepsis^11,12^. Mass spectrometry (MS)-based proteomics approaches have emerged as a particularly powerful technology for the comprehensive profiling of proteins comprising the plasma microenvironment^13^. For example, longitudinal profiling of postnatal proteomic changes has provided insights into the development of the immune system over the first weeks to months of life^14^. Untargeted proteomic analyses furthermore provide an unbiased approach to biomarkers discovery by removing the need to identify proteins of interest *a priori*.^15^

Proteomic profiling of neonatal cord blood provides a molecular snapshot at variable timepoints throughout neonatal development that could be used to reveal the underlying cellular processes occurring at birth, readiness for postnatal life, and for the identification of biomarkers specific to different disease states and prematurity-related complications.

While proteomic profiling of cord blood has demonstrated immunologic differences between preterm and term infants^16^, prior research has lacked inclusion of preterm infants across the continuum of gestational age and consideration of key perinatal characteristics such as the route of delivery, preeclampsia, intraamniotic infection, and neonatal sepsis that are likely to affect protein abundance. In this study, we have comprehensively characterized the cord blood proteome from infants born between 25 to 42 weeks using MS to provide a benchmark of normative cord blood proteomic profile and examine proteome differences across the developmental range of gestational ages.

## Methods

### Study cohort and specimen collection

We utilized archived cord blood plasma from an ongoing prospective study of infants born at Northwestern Prentice Women’s Hospital between 2008-2019. Parents were consented prior to or after birth; cord blood was centrifuged at 3000 rpm for 10 minutes and was separated into aliquots stored at -80 degrees Celsius until use. Samples in this investigation were selected from the biorepository based on gestational age and the absence of presumed or proven early onset neonatal sepsis (i.e., the infant received no antibiotic treatment course for sepsis within the first 72 hours of life and had no positive microbiologic sterile site cultures). A total of 150 infants were frequency matched within each gestational age (GA) category (epochs: 25-28 weeks, 29-32 weeks, 33-36 weeks, 37-42 weeks) with approximately equal numbers by sex, route of delivery (vaginal delivery vs. caesarean delivery with or without labor), and reason for preterm birth (maternal indication such as preeclampsia vs. fetal/pregnancy indication such as spontaneous preterm labor or preterm premature rupture of membranes). Clinical data including birth weight and intraamniotic infection were collected from the electronic medical record. This study was approved by the Institutional Review Boards of Northwestern University (STU00201858) and Lurie Children’s Hospital (IRB 2018-2145). Parental informed consent was obtained for use of clinical data and infant cord blood samples. All research activities were performed in accordance with the Declaration of Helsinki.

### Mass spectrometry sample preparation and analysis

Samples were thawed on ice and 20µl of plasma was utilized for study. Protein concentrations were determined using the Bicinchoninic Acid (BCA) method; untargeted mass spectrometry-based proteomic analysis was applied to 600 µg of extracted protein from each plasma sample. Samples were first depleted of fourteen known highly abundant proteins (Albumin, IgA, IgD, IgE, IgG, IgG (Light chains), IgM, Alpha-1-acid glycoprotein, Alpha-1-antitrypsin, Alpha-2-macroglobulin, Apolipoprotein A1, Fibrinogen, Haptoglobin, and Transferrin) using the Top 14 Abundant Protein Depletion Spin Columns (Thermo Scientific, Rockford, IL, USA). Remaining proteins were purified by acetone/TCA precipitation, reduced, alkylated, and digested with trypsin. Digested peptides were desalted on C18 columns (Thermo Scientific, Rockford, IL, USA) and eluted in 80% acetonitrile in 0.1% formic acid. Peptides were reconstituted with 0.1% formic acid in water and injected onto the in-house C18 trap column (3 cm length, 150 μm inner diameter, 3 μm particle size) coupled with an analytical C18 column (10.5 cm length, 75 μm inner diameter, 2 μm particle size, PicoChip).

Samples were separated using a linear gradient from 5% ACN/0.1% formic acid to 40% ACN/0.1% formic acid over 120 minutes using an UltiMate 3000 Rapid Separation nanoLC coupled to a Orbitrap Elite Mass Spectrometer (Thermo Fisher Scientific Inc, San Jose, CA). The full scans were acquired from 400-2000m/z at 60,000 resolving power and automatic gain control (AGC) set to 1×10^6^. The top fifteen most abundant precursor ions in each full scan were selected for fragmentation. Precursors were selected with an isolation width of 1 Da and fragmented by collision-induced dissociation (CID) at 35% normalized collision energy. Previously selected ions were dynamically excluded from re-selection for 58 seconds.

Samples were analyzed in duplicate, in a specified run order, across four batches. Samples were randomly assigned to batches using a stratified sampling approach to achieve balance on gestational age and other clinical characteristics (sex, type of delivery). A representative “pooled control,” including samples representing the full spectrum of the cohort, was used as an “internal standard” and run multiple times in each batch. Within each batch, the run order for samples and controls was determined by simple random sampling. MS raw files were analyzed with MaxQuant software (version 1.6.0.16).^17^ MS/MS-based peptide identification was carried out against the SwissProt human database with the Andromeda search engine in MaxQuant^18^ using a target-decoy approach to identify peptides and proteins at an FDR <1%. For LFQ, the MaxLFQ algorithm was used as part of the MaxQuant environment.^19^ The following modifications were set as search parameters: trypsin digestion cleavage after K or R (except when followed by P), 2 allowed missed cleavage sites, carbamidomethylated cysteine (static modification), and oxidized methionine, protein N-term acetylation (variable modification). Search results were validated with peptide and protein FDR, both at 0.01. Transformed (log_2_) LFQ values were used for all statistical analyses. The mass spectrometry proteomics data have been deposited to the ProteomeXchange Consortium via the PRIDE partner repository with the dataset identifier PXD051974.

### Proteomics data normalization

Boxplots representing the median and first and third quartiles were used to visualize the distribution of protein concentration among all proteins in pooled controls and identify the presence of any batch effects. To correct for batch effects demonstrated, batch normalization was conducted as follows.

Proteins that were detected in only one batch were excluded. Using the pooled control samples, the average difference in log_2_ LFQ value relative to the first batch was estimated using a linear regression model. Briefly, a beta coefficient for each batch was estimated using linear regression with batch one serving as the referent. The average protein difference from the first batch was then subtracted from the log_2_ LFQ value of each batch to determine the normalized log_2_ LFQ value.

Visual inspection of post-normalization protein levels by batch was used to determine the adequacy of the normalization procedure. The batch normalized protein abundance for each sample was averaged across each technical replicate for subsequent inter-patient analyses. If a protein was only detected in one of the replicates, the value of the batch normalized detected protein from the single replicate was used for analyses.

### Differential protein abundance determination

Separate linear regression models were used to examine the association between protein abundance and GA (unadjusted and adjusted for sex, labor, route of delivery, and preeclampsia). The response variable for each model was the batch normalized value log_2_ transformed protein level for the given protein. Proteins were included in adjusted models if found in more than one sex, delivery category, and preeclampsia category. The primary explanatory variable of interest was GA. Scatter plots were examined to determine whether GA demonstrated a linear or nonlinear association (e.g., using splines or quadratic terms) with protein level. The relationships between GA and protein abundance, in general, appeared linear across proteins, so a linear term for GA was included in models. There were seven sets of twins among the 150 controls. One twin from a pair was randomly selected to be included in the models (n=143). To control Type 1 error rate, P-values were adjusted for multiple testing using the Benjamini-Hochberg False Discovery Rate (FDR) method, and associations with FDR-adjusted P values <0.05 were considered statistically significant^20^.

### Functional enrichment and visualization

The relative expression abundance of all proteins that changes significantly over gestational age was visualized in a heat map. The batch normalized protein values were z-score normalized by subtracting the relative protein abundance within a given specimen by the mean abundance across all specimens in which the protein was detected and then dividing by standard deviation. Proteins that were undetected in more than 50% of specimens were excluded from visualization. Z-score normalized values were visualized in a heatmap using the *clustermap* function in the *seaborn* (v 0.11.1) package within the python (v 3.8.8) environment with specimens ordered left to right by GA and proteins clustered by z-score profile from top to bottom. The *clustermap* function uses hierarchical clustering with average linkage and Euclidean distance.

Functional enrichment analysis of the proteins found to be significantly increased or decreased in abundance was performed using MetaScape v3.5 (https://metascape.org/gp/index.html#/main/step1)^21^. UniProt IDs were used as unique identifiers; two isoforms of APOB (P04114), PLG (P00747), and FGA (P02671) were consolidated and immunoglobulins were excluded (P0DOX5, P0DOX7, P0DOY3, and P01859) for final analysis of 64 proteins. All proteins detected in the overall proteomic dataset (n = 465) were set as the background gene set before enrichment. Protein-protein interaction networks were visualized using STRING v11.5 (https://string-db.org/)^22^. Network visualization was limited to physical subnetworks based on experiment and database active interaction sources with a 0.15 minimum interaction score required. Nodes were colored by an increase or decrease in abundance with edge width reflective of protein interaction confidence score. Proteins contributing to significantly enriched pathways were annotated with colored boxes.

## Results

### Patient demographics

The distribution of GA and associated clinical/demographic details for the 150 infants included in this study are displayed in **Table 1**. The mean GA across all infants was 33.2 weeks (standard deviation 4.5, range 25.9-41.4). 17 infants (11%) were 25-28 weeks, 43 (29%) were 29-32 weeks, 50 (33%) were 33-36 weeks, and 40 (27%) were 37 weeks and greater. 77 (51%) of the infants were female. 34 (23%) infants were born to women with preeclampsia. 44 (29%) infants were from 22 individuals with multiple gestations, all of whom were born at less than 37 weeks GA. Only one infant was born to an individual who had clinical chorioamnionitis.

**Table 1.** Demographics and clinical covariates.

| **n=150** | **Patients**<br>**n (%)** |
| --- | --- |
| **Gestational age** weeks<br>median (IQR) | 33.7 (29.6-37.5) |
| 25-28 | 17 (11%) |
| 29-32 | 43 (29%) |
| 33-36 | 50 (33%) |
| ≥37 | 40 (27%) |
| **Infant sex (female)** | 77 (51%) |
| **Labor and delivery** |  |
| vaginal with labor | 77 (51%) |
| caesarean with labor | 42 (28%) |
| caesarean without labor | 31 (21%) |
| **Preeclampsia** | 34 (23%) |
| **Multiple gestation** | 44 (29%) |
| **Clinical chorioamnionitis** | 1 (0.7%) |

### Differential protein abundance across GA

The total BCA, representative of protein abundance, is positively correlated with GA (**Figure 1**). Of the 465 unique proteins identified in control plasma samples, 391 were included in the adjusted regression models (adjusted for sex, preeclampsia, and delivery route). Proteins were excluded from adjusted multivariable regression models if they were only found in one group of the covariate categories (for example found in only male infants). Gestational age was associated with protein abundance in 70 proteins with FDR-adjusted P-value <0.05 (**Supplemental Table 1**). To visualize each protein’s change over GA, the normalized protein abundance in each specimen was plotted relative to GA of the infant (**Figure 2**). The slope (‘beta’ value) of the linear fitted model for each protein is provided in **Supplemental Table 1.** Representative plots in **Figure 2** demonstrate examples of proteins with positive (e.g., plasminogen; **Figure 2A**) and negative (e.g., alpha-fetoprotein; **Figure 2B**) correlation between protein abundance and GA. These changes are summarized in a volcano plot (**Figure 3**) depicting the log_10_ of the FDR-adjusted P-values and the associated betas from linear regression models. Proteins such as alpha-fetoprotein, collagen alpha-1(V) chain, and basement membrane-specific heparan sulfate proteoglycan core protein are highly abundant earlier in gestational development while many immunologically active proteins are more abundant later, including IgG-1 chain C region, complement C1q subunit C, and protein S100-A9, a calcium- and zinc-binding protein which plays a prominent role in the regulation of inflammatory response (all aforementioned proteins with p<0.0001).

**Figure 1.**
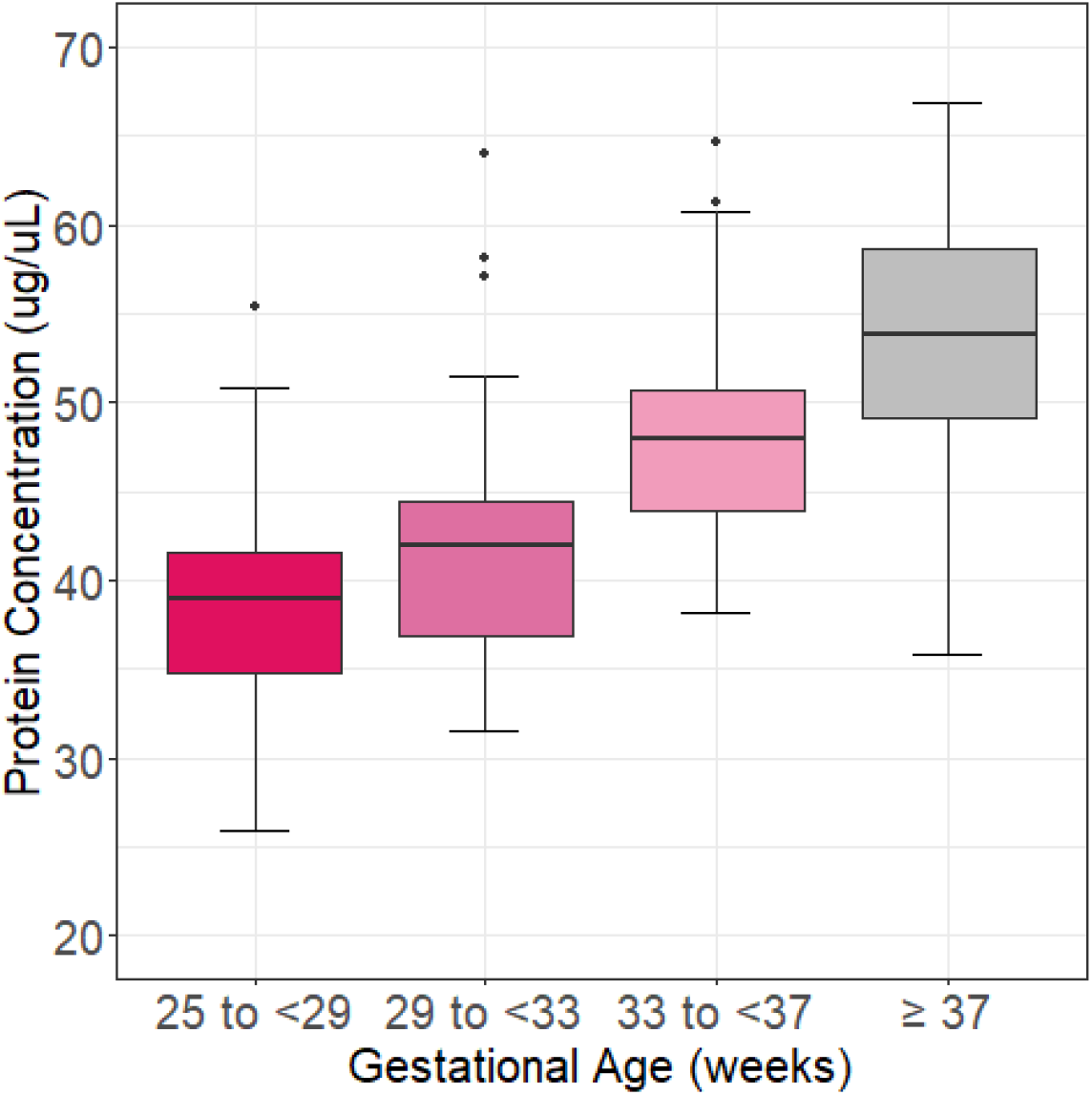
Total protein concentration in each plasma sample. Box and whisker plot of protein concentration (ug/uL) distribution across gestational age (GA) categories. The lower and upper ends of each box correspond to the 25th and 75^th^ percentiles for a given group [shaded area is the interquartile range (IQR)]. The black line in each box is the median. The whiskers represent the largest and smallest observed data points that are no further than +/-1.5 times the IQR, respectively. Points outside of the boundary of the whiskers are outliers. Kruskal-Wallis across GA categories p<0.0001.

**Figure 2.**
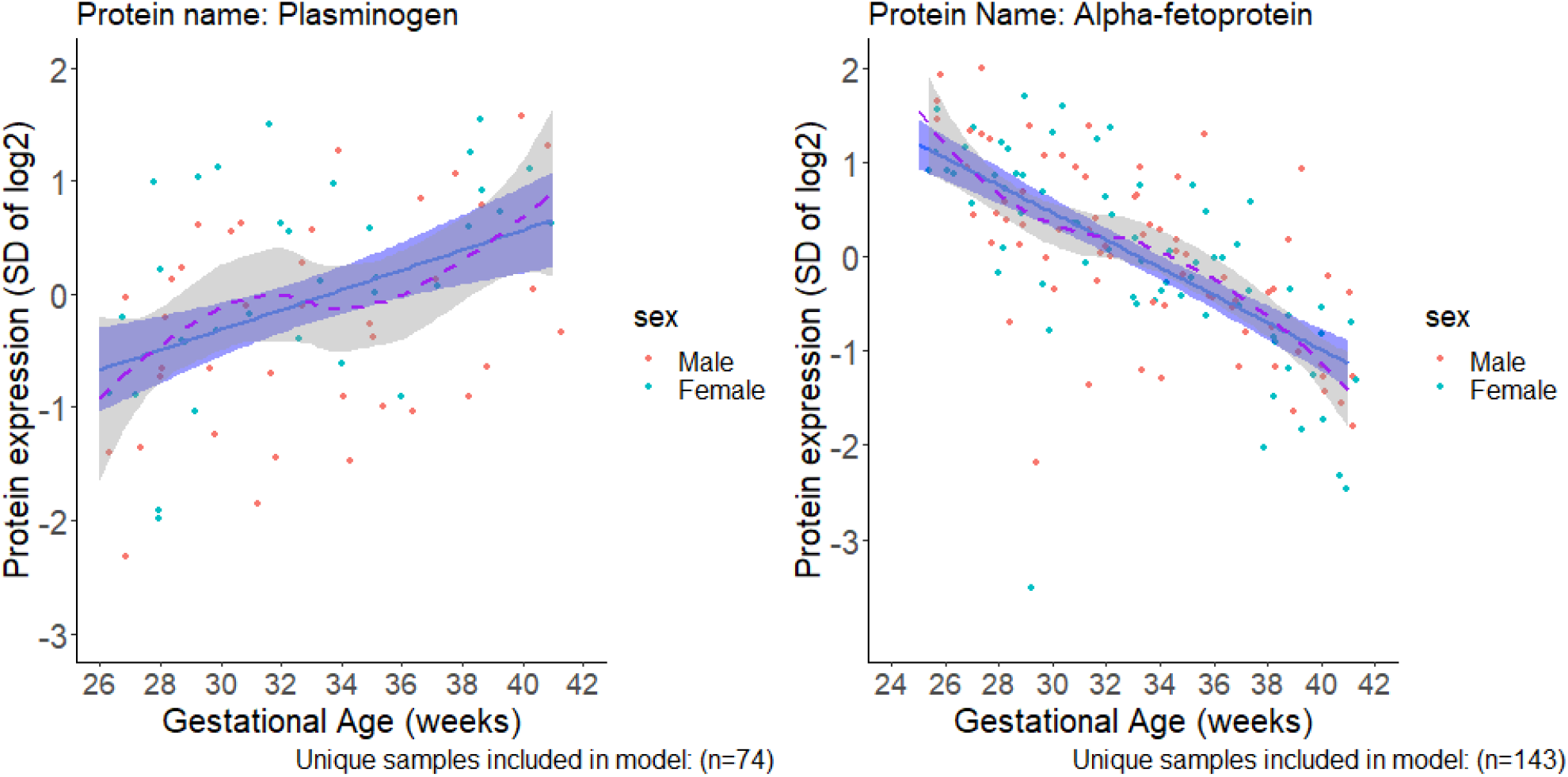
Representative plots of relative protein abundance by gestational age. The scatter plots show the observed values by sex of newborn. The blue line and shaded blue show the fitted linear model and 95% confidence interval (CI) of the association respectively. The purple dashed line shows a loess smoothed line of the association, and the 95% CI is the shaded gray region (most appeared approximately linear). **A)** Plasminogen model included n=74 samples. **B)** Alpha-fetoprotein included n=143 unique samples in the model.

**Figure 3.**
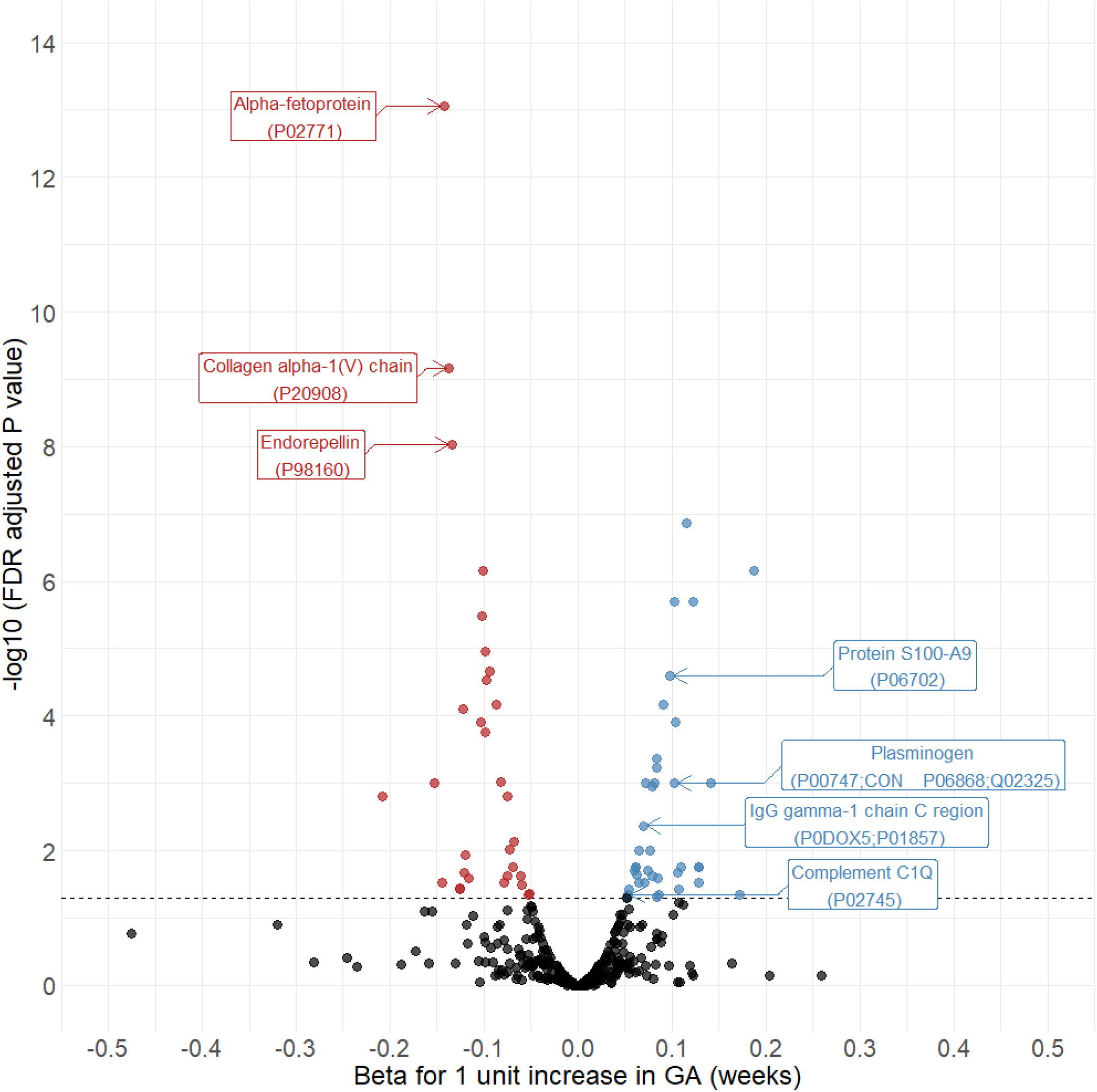
Volcano plot of protein abundance association with gestational age. Shown are -log_10_ of the FDR adjusted P values and betas from linear regression models for a unit increase in continuous gestational age term on a standard deviation increase in protein abundance for a given protein adjusted for sex, preeclampsia, labor route of delivery. Colors show direction of linear associations (positive [blue] indicates increasing GA associated with increasing protein abundance and negative [red] indicates decreasing GA associated with decreasing protein abundance). Seventy proteins were found to be significantly associated with gestational age in adjusted models. Proteins with FDR adjusted p <0.0001 are labelled. The full list of proteins with FDR adjusted p<0.05 can be found in Supplementary Table 1.

### Visualization and pathway analysis

To better visualize the differences in protein levels across GA, we plotted a heatmap of the normalized protein levels for each significantly changing protein identified above in each specimen ordered along the x-axis by GA (**Figure 4A**). Proteins without detectable levels in more than 50% of specimens (n = 15) were excluded from visualization and hierarchical clustering was used to group proteins by similarity in abundance trends over GA. This highlights several distinct groups of proteins where levels change over time. For example, COL5A1, CD14, HSPG2, QSOX1, FCGBP seem to be abundant in early GA, but decrease as GA increases. This trend is also apparent in the cluster located at the bottom half of the heatmap that includes CD109, COL1AI, APOC3, APOE, TGFBI, AFP, AGT, APOB, LUM, SERPINA1, B2M, FGA, THBS4, F13A1 and SERPINA5. However, several proteins also follow the opposite trend with lower abundance early and higher abundance late, including HBA1, HBB, HPX, IGFALS, CP, AFM, SERPINF2, SERPIND1, A2M, ATRN, PGLYRP2, IGHG1, C7, ITIH1, PLG, F2, SERPINC1, C1QC and C1QA.

**Figure 4.**
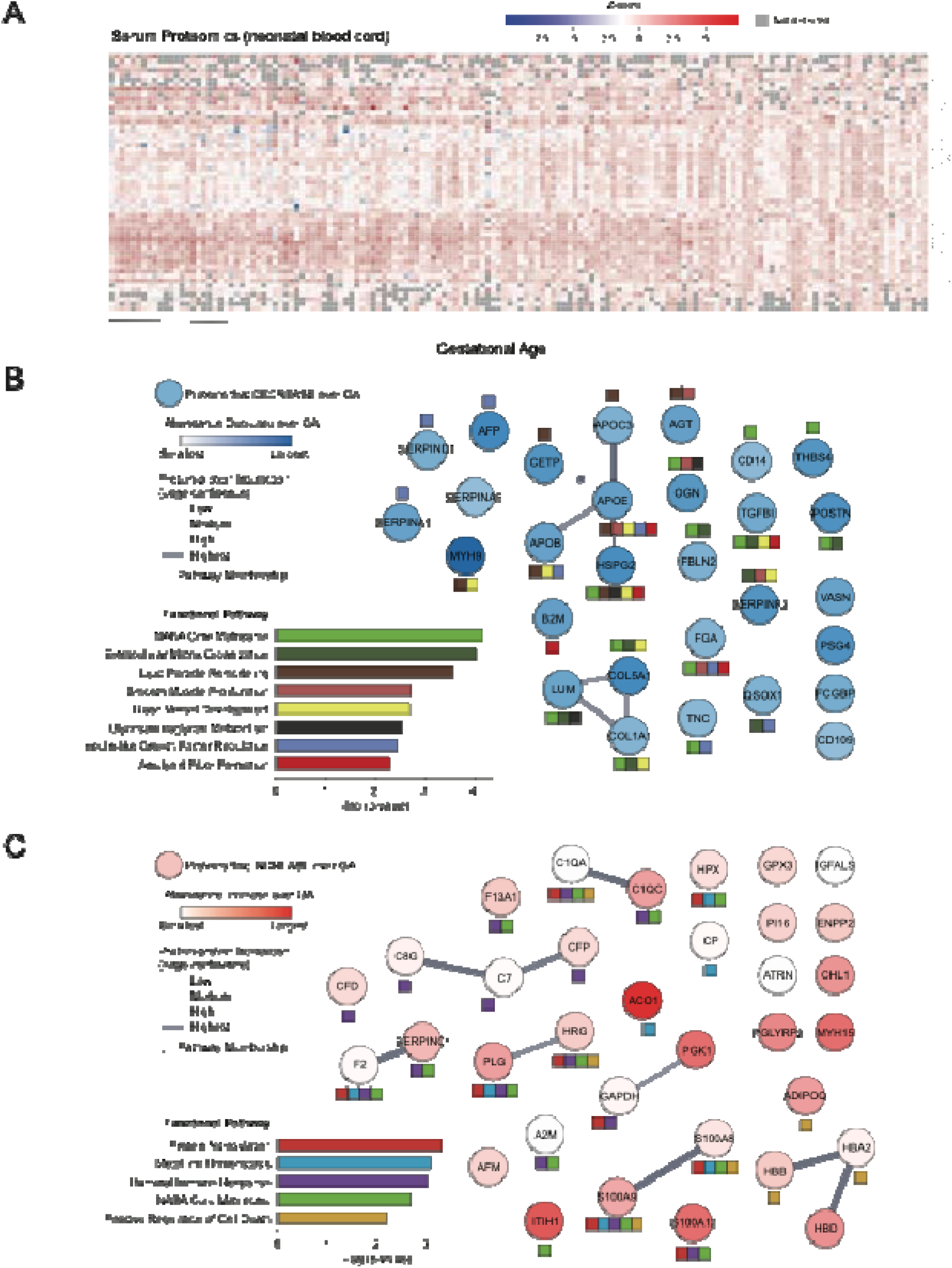
Functional enrichment analysis of proteins in neonatal cord blood that change over gestational age. **A)** Heatmap of the protein Z-scores detected in neonatal cord blood samples arranged by gestational age from left to right. Proteins are grouped top to bottom by hierarchical clustering. Functional enrichment analysis and protein-protein interaction networks of proteins significantly **B)** decreased or **C)** increased over gestational age are shown below. Network nodes are shaded by abundance change over gestational age with edge width reflecting protein-protein interaction confidence. Significantly enriched pathways are highlighted in colored bar charts to the left; each protein that maps to the identified pathways is indicated by a color-matched box beneath the network node.

Functional enrichment analysis was performed for those proteins found to be significantly decreased in relative abundance (n = 29, **Figure 4B**) or increased in abundance (n = 34, **Figure 4C**) with increasing GA (7 identifiers did not map back to unique proteins and were excluded from analysis, see **Methods**). Proteins that decreased in abundance were enriched for eight pathways: NABA core matrisome, extracellular matrix organization, lipid particle remodeling, smooth muscle proliferation, blood vessel development, glycosaminoglycan metabolism, insulin-like growth factor regulation, and amyloid fiber formation. These enriched protein sets include several components of known protein complexes. For example, the proteoglycan LUM and the collagen proteins COL5A1 and COL1AI form high confidence protein interactions and are all associated with extracellular matrix organization pathways. Likewise, CETP, APOC3, APOB, and APOE are known to interact and play critical roles in lipid particle remodeling. Several factors implicated in insulin-like growth factor regulation were also decreased, including SERPINF2, SERPIND1, FGA, and SERPINA5.

More proteins were found to increase in abundance over GA than decrease. However, these were associated with a narrower set of pathways, specifically: protein nitrosylation, metal ion homeostasis, humoral immune response, NABA core matrisome, and positive regulation of cell death. One well-known transition that occurs in the serum throughout development is the swapping of hemoglobin subunits from γ-globin in neonates to β-globin and δ-globin gene expression in pediatric and adult patients^23^. Consistent with this transition, we see increased abundance of β-globin (HBB) and δ-globin (HBD), as well as α-globin (HBA2). We additionally see the increased abundance of several proteins associated with the immune response, including several complement proteins (C1QA, C1QC, C7, CFP, and C8G) and several S100 calcium binding proteins (S100A9, S100A12, and S100A8). Notably, several immunoglobulins were also increased over the course of GA (specifically kappa light chain, lambda light chain, and gamma heavy chains), though these are not visualized here.

## Discussion

Our data demonstrate that the abundance of several cord blood proteins varies significantly across GA. Proteins that function in structural development and growth, including extracellular matrix organization, lipid particle remodeling, blood vessel development, and insulin-like growth factor regulation, are more abundant earlier in gestation. Later in gestation, proteins involved in immune response pathways, including complements, and calcium-binding proteins involved in inflammation are higher in abundance. These data highlight the differences in immunologic state across GA and provide insights into the higher risk of invasive infections among preterm infants. Furthermore, these data contribute to the knowledge of the physiologic state of neonates across GA, which is crucial to understand as we: 1) strive to emulate the *in utero* environment to best support the developmental process of those born preterm, 2) understand mechanisms of pathology that cause adverse health outcomes for preterm infants, and 3) develop cord blood markers for neonatal disease conditions that can predict and help tailor medical management.

In a 2021 review of proteomic studies that attempted to identify biomarkers for prematurity-related diseases, Letunica *et al*. determined that only 13% of studies investigated cord blood even though cord blood is a readily available specimen at birth.^11^ Suski *et al.* investigated the cord blood proteome of preterm infants in three GA groups (<=26 weeks, 27-28 weeks, and 29-30 weeks) and compared them to the proteomes of a full term control group. They reported differences in inflammatory, immunomodulation, coagulation, and complement systems in preterm versus term infants.^16^ Specifically, they found that preterm infants had decreased levels of anti-inflammatory proteins (e.g., orsomucoid isoforms) and B-cell mediated immunity markers, and increased abundance of inflammatory proteins such as leucine-rich alpha-2-glycoprotein (LRG1) and complement activation cascades, a finding that complements our results of lower proteins related to humoral immunity at earlier GA.

However, the authors also suggest an increase in inflammatory mediators in preterm infants, whereas out results showed increased inflammatory and immune response proteins and complement components later in gestational age. Our pathway analysis revealed many of the proteins that function in inflammatory signaling and immune response are lower in preterm infants with no infection. A likely explanation for this notable difference is that our research excluded preterm infants with early onset sepsis, and thus represents the state of the cord blood proteome in the absence of infection. Given that treatment of early-onset sepsis is common in very preterm infants, the analysis of cord blood inflammatory proteins may be skewed if one has not accounted for infection.

Other types of immune phenotyping have been reported in cord blood across GA. Olin *et al.* noted differences in both cord blood proteins and decreased neutrophil proportions in preterm compared to term infants. They reported an increase in inflammatory cord blood proteins, attributed to the role of inflammation and infection in preterm birth, but not reflecting gestational norms without infection.^14^ Anderson *et al.* utilized flow cytometry and cytokine assays of cord blood to compare preterm infants (30-34 weeks GA) to full-term infants.^24^ They found that preterm infants had lower frequencies of monocytes, NK cells, CD8+ T-cells and gamma-delta T-cells than their term full term counterparts. There were increased intermediate monocytes, CD4 T cells, Tregs, and transitional B-cells in preterm infants indicating immaturity of the innate immune system and a skewed cellular landscape related to increased susceptibility of preterm infants to bacterial and viral infections. They also noted lower levels of pro-inflammatory cytokines and chemokines in preterm infants, further confirming preterm infants impaired ability to fight off infection. Finally, Peterson *et al.* applied single-cell immunoprofiling of cord blood for 45 infants (20 preterm) after excluding infants exposed to clinical chorioamnionitis or with active infection.^25^ The study also controlled for potential other clinical confounders, including steroid administration. They found a strong relationship between GA and the neonatal immune profile at birth. Specifically, increasing GA was associated with a progressive increase in the ligand-specific responsiveness to immune system stimulation. This finding aligns with our finding of increased cell-signaling, calcium binding, and immune response proteins with later gestational age. Our work supports the conclusion that decreased antigen- and cytokine-specific immune responses may contribute to preterm infant susceptibility to infection.

Furthermore, differences in proteins across GA may provide insight into underlying pathophysiology and risk of pathology. Functional analysis identified several pathways associated with increased abundance of proteins that are implicated in vascular development, lipid metabolism, smooth muscle proliferation, insulin-like growth factor regulation, and the matrisome. For example, afamin, an anti-inflammatory protein previously hypothesized to be a hallmark of detrimental oxidative stress and related to retinopathy of prematurity, is less abundant in the cord blood of preterm infants.^16^ The process of in utero development represents a complex and dynamic system between the pregnant person and fetus. Through this study of neonates born from 25-42 weeks GA, we aim to help establish the baseline state of the developmental continuum.

The strengths of this study include: 1) the analysis of cord blood proteomics on a large sample size across the GA spectrum; 2) precise clinical categorization and consideration of covariates that may impact the cord blood proteome including exclusion of infants with early onset infection and adjustment for labor, preeclampsia, and sex; and 3) careful methodologic and data normalization, both in design and analysis of discovery mass spectrometry proteomics (distribution and normalization across batches, pooled control, addressing missingness). Additionally, functional pathway analysis strengthens our ability to parse key pathways of relevance and provide validation through the demonstration of known GA-related differences in hemoglobin and immunoglobulin proteins^26^. Limitations include that mass spectrometry proteomics does not provide absolute quantitation of protein, but rather spectral counts and relative abundance. The detectable protein abundance reflects the level after potential clearance, degradation, or transport/localization of expressed proteins to compartments. For this reason, we highlight relative abundance and levels of proteins rather than using terms akin to protein expression (i.e., “up/down-regulation”). Thus, specific biomarker development warrants quantitative validation methods. Additionally, the cord blood specimen used in this analysis was intended to be obtained at the time of birth from the umbilical vein. However, it is possible that there is some mixing of umbilical arterial and venous blood. Prior literature raises questions about mediating cord blood markers by placental clearance and whether cord blood proteins may reflect maternal serum. In multiple studies, paired analysis of maternal and fetal cord blood biomarkers has shown weak or no correlation.^27,28^

In conclusion, our study utilizing untargeted proteomics has demonstrated that the cord blood proteome varies significantly with GA at birth. There are meaningful differences in several pathways, including crucial aspects of inflammation and immune response. Future research can apply this knowledge of the baseline state to find methods to develop more precise, GA-specific cord blood diagnostic markers of short and perhaps long-term^29^ health and disease.

## Data Availability

The mass spectrometry proteomics data have been deposited to the ProteomeXchange Consortium via the PRIDE partner repository with the dataset identifier PXD051974.

## Acknowledgments

The authors would like to acknowledge Erin Cullather, BS; Paul Martin Thomas, PhD; Aaron Hamvas, MD; and Thomas Shanley, MD for their support toward this work. Proteomics services were performed by the Northwestern Proteomics Core Facility, generously supported by NCI CCSG P30 CA060553 awarded to the Robert H Lurie Comprehensive Cancer Center, instrumentation award (S10OD025194) from NIH Office of Director, and the National Resource for Translational and Developmental Proteomics supported by P41 GM108569.

## Author Contributions

LBM: conceptualization, methodology, data curation, analysis, funding acquisition, writing - original draft. B-KC and YG: methodology, investigation, data curation, writing - review & editing. DS, NL, TL-H and JFH: methodology, data curation, formal analysis, visualization, writing - review & editing. SO and NJR: methodology, investigation, writing - review & editing. WAG: conceptualization, methodology, supervision, writing - review & editing. KM: conceptualization, methodology, data curation, supervision, writing - review & editing. PCS: conceptualization, methodology, data curation, supervision, writing - review & editing, funding acquisition. All authors contributed to the article and approved the submitted version.

## Data Availability Statement

The original contributions presented in the study are included in the article/supplementary material, further inquiries can be directed to the corresponding author/s. The raw data supporting the conclusions of this article will be made available by the authors, without undue reservation and is also available on Proteome Xchange Consortium via the PRIDE partner repository (dataset PXD051974).

## Ethics Statement

Northwestern University and Lurie Children’s Institutional Review Boards reviewed and approved the studies involving human participants (Northwestern STU00201858, Lurie IRB 2018-2145). The patients/participants provided their written informed consent to participate in this study.

