## Supplemental Table for "Evolution of the Umbilical Cord Blood Proteome Across Gestational Development"

### Supplemental Table of Significantly Varied Proteins

| Uniprot ID | Protein Names | Beta (SE) | P value | Adjusted P value |
|---|---|---|---|---|
| P02771 | Alpha-fetoprotein | -0.142 (0.015) | <0.001 | <0.001 |
| P20908 | Collagen alpha-1(V) chain | -0.137 (0.018) | <0.001 | <0.001 |
| P98160 | Endorepellin | -0.134 (0.019) | <0.001 | <0.001 |
| Q96PD5;CON__ENSEMBL:ENSBTAP00000016285 | N-acetylmuramoyl-L-alanine amidase | 0.115 (0.018) | <0.001 | <0.001 |
| P01019 | Angiotensinogen | -0.101 (0.017) | <0.001 | <0.001 |
| P0DOX7 | | 0.187 (0.024) | <0.001 | <0.001 |
| P01859 | Ig gamma-2 chain C region | 0.122 (0.021) | <0.001 | <0.001 |
| P02747 | Complement C1q subcomponent subunit C | 0.103 (0.018) | <0.001 | <0.001 |
| P61769 | Beta-2-microglobulin | -0.102 (0.018) | <0.001 | <0.001 |
| P01009 | Alpha-1-antitrypsin | -0.099 (0.018) | <0.001 | <0.001 |
| P51884;CON__Q05443 | Lumican | -0.094 (0.018) | <0.001 | <0.001 |
| P06702 | Protein S100-A9 | 0.098 (0.019) | <0.001 | <0.001 |
| Q6EMK4 | Vasorin | -0.098 (0.019) | <0.001 | <0.001 |
| P01008 | Antithrombin-III | 0.091 (0.019) | <0.001 | <0.001 |
| Q15582 | Transforming growth factor-beta-induced protein ig-h3 | -0.087 (0.018) | <0.001 | <0.001 |
| P11597 | Cholesteryl ester transfer protein | -0.122 (0.024) | <0.001 | <0.001 |
| P02649;CON__Q03247 | Apolipoprotein E | -0.103 (0.021) | <0.001 | <0.001 |
| Q15848 | Adiponectin | 0.103 (0.022) | <0.001 | <0.001 |
| P04114;CON__ENSEMBL:ENSBTAP00000032840 | Apolipoprotein B | -0.099 (0.021) | <0.001 | <0.001 |
| P68871 | Hemoglobin subunit beta | 0.083 (0.019) | <0.001 | <0.001 |
| P00488 | Coagulation factor XIII A chain | 0.083 (0.019) | <0.001 | <0.001 |
| Q9Y6R7 | IgG Fc-binding protein | -0.083 (0.020) | <0.001 | <0.001 |

### Supplemental Table of Significantly Varied Proteins

| Uniprot ID | Protein Names | Beta (SE) | P value | Adjusted P value |
|---|---|---|---|---|
| P00747;CON__P06868;Q02325 | Plasminogen | 0.103 (0.024) | <0.001 | 0.001 |
| P02790 | Hemopexin | 0.072 (0.017) | <0.001 | 0.001 |
| P04196 | Histidine-rich glycoprotein | 0.081 (0.020) | <0.001 | 0.001 |
| P19827 | Inter-alpha-trypsin inhibitor heavy chain H1 | 0.141 (0.030) | <0.001 | 0.001 |
| P04114 | Apolipoprotein B | -0.153 (0.033) | <0.001 | 0.001 |
| P43652;CON__REFSEQ:XP_585019 | Afamin | 0.079 (0.019) | <0.001 | 0.001 |
| P02656 | Apolipoprotein C-III | -0.075 (0.019) | <0.001 | 0.002 |
| P35579;P35580;Q7Z406 | Myosin-9 | -0.208 (0.034) | <0.001 | 0.002 |
| P0DOX5;P01857 | Ig gamma-1 chain C region | 0.070 (0.019) | <0.001 | 0.004 |
| P02452 | Collagen alpha-1(I) chain | -0.068 (0.019) | <0.001 | 0.007 |
| O00391 | Sulfhydryl oxidase 1 | -0.073 (0.021) | <0.001 | 0.010 |
| P22352 | Glutathione peroxidase 3 | 0.076 (0.022) | <0.001 | 0.010 |
| P69905 | Hemoglobin subunit alpha | 0.065 (0.019) | <0.001 | 0.010 |
| P35443 | Thrombospondin-4 | -0.120 (0.033) | 0.001 | 0.012 |
| P00450 | Ceruloplasmin | 0.061 (0.019) | 0.002 | 0.018 |
| P02671;CON__P02672 | Fibrinogen alpha chain | -0.069 (0.022) | 0.002 | 0.018 |
| Q9Y2K3 | Myosin-15 | 0.128 (0.037) | 0.002 | 0.018 |
| P00734;CON__P00735 | Prothrombin | 0.061 (0.019) | 0.002 | 0.018 |
| P02042;CON__Q3SX09 | Hemoglobin subunit delta | 0.110 (0.032) | 0.002 | 0.018 |
| P80511 | Protein S100-A12; Calcitermin | 0.128 (0.039) | 0.002 | 0.018 |
| P00746 | Complement factor D | 0.075 (0.024) | 0.002 | 0.020 |
| P10643 | Complement component C7 | 0.061 (0.019) | 0.002 | 0.020 |

### Supplemental Table of Significantly Varied Proteins

| Uniprot ID | Protein Names | Beta (SE) | P value | Adjusted P value |
|---|---|---|---|---|
| P00747;CON__P06868;Q15195;Q02325 | Plasminogen | 0.106 (0.032) | 0.002 | 0.021 |
| P08697 | Alpha-2-antiplasmin | -0.121 (0.037) | 0.002 | 0.021 |
| P04406 | Glyceraldehyde-3-phosphate dehydrogenase | 0.062 (0.020) | 0.003 | 0.023 |
| P24821 | Tenascin | -0.061 (0.020) | 0.003 | 0.024 |
| P98095 | Fibulin-2 | -0.075 (0.025) | 0.003 | 0.024 |
| Q6UXB8 | Peptidase inhibitor 16 | 0.079 (0.026) | 0.003 | 0.024 |
| P20774 | Mimecan | -0.116 (0.036) | 0.003 | 0.026 |
| P0DOY3;P0DOY2;P0DOX8;P0CF74;P0CG04;B9A064;A0M8Q6 | Immunoglobulin lambda-6 chain C region | 0.086 (0.028) | 0.003 | 0.026 |
| P00558;P07205 | Phosphoglycerate kinase 1 | 0.128 (0.040) | 0.004 | 0.030 |
| P05546 | Heparin cofactor 2 | -0.079 (0.027) | 0.004 | 0.030 |
| P07360 | Complement component C8 gamma chain | 0.065 (0.022) | 0.004 | 0.030 |
| P02671 | Fibrinogen alpha chain | -0.145 (0.047) | 0.004 | 0.030 |
| P05109 | Protein S100-A8 | 0.070 (0.024) | 0.004 | 0.030 |
| P08571 | Monocyte differentiation antigen CD14 | -0.060 (0.021) | 0.005 | 0.033 |
| Q15063;CON__Q2KJC7 | Periostin | -0.126 (0.042) | 0.005 | 0.036 |
| P35858 | Insulin-like growth factor-binding protein complex acid labile subunit | 0.055 (0.019) | 0.006 | 0.037 |
| O00533 | Neural cell adhesion molecule L1-like protein | 0.108 (0.036) | 0.006 | 0.037 |
| Q00888;Q15238 | Pregnancy-specific beta-1-glycoprotein 4/5 | -0.126 (0.041) | 0.006 | 0.037 |
| P05154 | Plasma serine protease inhibitor | -0.052 (0.019) | 0.007 | 0.044 |
| P02745 | Complement C1q subcomponent subunit A | 0.053 (0.020) | 0.007 | 0.045 |
| P27918 | Properdin | 0.086 (0.031) | 0.008 | 0.046 |

### Supplemental Table of Significantly Varied Proteins

| Uniprot ID | Protein Names | Beta (SE) | P value | Adjusted P value |
|---|---|---|---|---|
| P21399 | Cytoplasmic aconitate hydratase | 0.172 (0.057) | 0.008 | 0.046 |
| Q6YHK3 | CD109 antigen | -0.053 (0.020) | 0.008 | 0.046 |
| O75882 | Attractin | 0.053 (0.020) | 0.008 | 0.049 |
| Q13822 | Ectonucleotide pyrophosphatase/phosphodiesterase family member 2 | 0.083 (0.030) | 0.009 | 0.049 |
| P01023 | Alpha-2-macroglobulin | 0.053 (0.020) | 0.009 | 0.049 |

Table of proteins where gestational age had a significant linear association at FDR adjusted P value of <0.05. The Uniprot ID and Protein Name and beta coefficient (SE) from the linear model for the gestational age term adjusted for child sex, preeclampsia, and labor and delivery type are shown. The unadjusted and FDR adjusted P values are also included in the table.
